# Mechanism-Specific Differences in Traumatic Brain Injury: A Retrospective Urban Trauma Cohort Study

**DOI:** 10.64898/2025.12.28.25341910

**Authors:** Raymond Zhou, Shameeke Taylor

## Abstract

**Background:** Traumatic brain injury (TBI) remains a major cause of morbidity, yet U.S. comparisons of injury mechanisms in diverse urban settings are limited. We examined differences across motorized road injuries, non-motorized road injuries, and falls.

**Methods:** We performed a retrospective study of consecutive TBI admissions to an ACS-verified trauma center from 2017 to 2022. Mechanisms were categorized as motorized, non-motorized, or falls. Outcomes included mortality, ICU and ventilator use, and hospital length of stay (LOS). Secondary measures included GCS-based TBI severity and orthopedic injury. Group differences were evaluated using chi-square/Fisher exact and Kruskal-Wallis tests; motorized versus non-motorized road injuries were compared with Wilcoxon rank-sum tests.

**Results:** Among 1,131 TBI admissions, falls predominated (90.2%), followed by non-motorized (5.5%) and motorized (4.3%) injuries. Fall-related TBI occurred in older adults (mean age 71.2 vs 49.6 non-motorized and 45.6 motorized). Road injuries affected more frequently male (≈75 to 80% vs 61%; p=0.005) and Hispanic patients, who comprised 37.1% of non-motorized and 36.7% of motorized injuries versus 23.8% of falls. Mean Injury Severity Score (ISS) was 11.9 (non-motorized), 15.1 (motorized), and 14.1 (falls); ICU days were 1.7, 4.7, and 2.2; LOS 5.4, 8.4, and 8.3 respectively. Ventilator use differed across mechanisms (p=0.02), as did orthopedic injury (p=0.009). Mortality was highest after motorized injuries (14.3%) compared with falls (7.5%) and non-motorized injuries (3.2%). In pairwise comparisons, motorized injuries showed higher ISS, greater ICU and ventilator needs, and longer LOS (all p≈0.01 to 0.02), with a trend toward lower GCS (p=0.058).

**Conclusions:** Falls accounted for most TBIs and primarily affected older adults, whereas motorized road injuries, though less frequent, produced the greatest severity, resource utilization, and mortality. The elevated representation of Hispanic patients in road-related mechanisms highlights a need for targeted prevention in urban communities.

## Introduction

Traumatic brain injury (TBI) remains a major global health concern and a leading cause of death and disability. Recent analyses of the *Global Burden of Disease Study* estimate 20–27 million new cases annually, a figure that continues to rise due to population aging and evolving risk exposures.^1^□□ The incidence of TBI varies across demographic groups—men experience higher rates than women, with peak incidence among men aged 20–24 years, likely reflecting risk-taking behaviors.^1^ In contrast, adults aged ≥75 years have the highest rates of TBI-related hospitalization and death in the United States, often due to falls, comorbidities, and medication-related complications.□□□ These demographic patterns underscore the need to better understand mechanisms of injury as potential mediators of TBI risk and outcome.

Mechanisms of TBI differ by population and may confound observed associations with severity and prognosis. Falls and road injuries account for most TBIs worldwide^1^□□ and in the United States,□□□ though their relative contributions differ by region and income level. Road traffic injuries predominate in low- and middle-income countries, whereas falls now exceed road injuries in high-income nations, particularly among older adults.^1^□□ In the US, men more frequently sustain TBIs from motor vehicle crashes, self-harm, and assault, while women experience more fall-related TBIs.□ Prior international studies, including the *International Traumatic Coma Project*, have shown that road-related TBIs are typically more severe— associated with higher Injury Severity Scores (ISS), lower Glasgow Coma Scores (GCS), and longer intensive care unit (ICU) and ventilation durations—yet paradoxically result in better survival and functional recovery than fall-related injuries.^9-10^ These findings highlight the complex relationship between injury mechanism, clinical course, and outcome.

Further stratification of road-related TBIs into motorized (e.g., automobiles, motorcycles, e-bikes) and non-motorized mechanisms (e.g., bicycles, scooters) reveals distinct demographic and clinical patterns. Studies from Europe have shown that motorcyclists and motorists sustain more severe injuries, reflected by lower GCS and higher intracranial hemorrhage rates, compared to bicyclists who are typically older and experience more favorable outcomes.^9-12^ However, most comparative studies have been conducted outside the United States, limiting generalizability to its diverse urban populations, where transportation patterns, alcohol use, and healthcare access differ substantially. Moreover, factors such as insurance status, alcohol use (EtOH), and polytrauma have been associated with morbidity and mortality, complicating interpretation of injury severity and outcomes.^13-^

Despite increasing attention to global TBI epidemiology, few comparative studies of TBI mechanisms have been conducted in the United States, where demographic diversity and healthcare systems differ markedly from those of prior international cohorts. We therefore sought to examine the epidemiology, risk factors, and outcomes of TBI by mechanism— motorized vehicle, non-motorized device, and falls—in a large, diverse, metropolitan population. We hypothesized that (1) motorized vehicles would be associated with the most severe injuries as measured by GCS, ISS, and hospital course, and (2) that both motorized and non-motorized mechanisms would predominantly affect younger patients relative to falls. Given the rapid proliferation of micromobility devices in urban environments like New York City, these findings have important implications for clinical management, injury prevention, and public health policy.

## Methods

We conducted a retrospective cohort study of consecutive trauma admissions at an urban, American College of Surgeons–verified Level I trauma center between 2017 and 2022. The institutional trauma registry, formatted according to the National Trauma Registry of the American College of Surgeons (NTRACS) standards, was queried to identify patients with traumatic brain injury (TBI). Eligible patients included those with a traumatic mechanism and TBI documented by registrar abstraction, defined as a head Abbreviated Injury Scale (AIS) score greater than zero and/or a clinical or imaging diagnosis of TBI, who were admitted to or observed in the hospital. Patients with non-traumatic intracranial conditions or unknown mechanisms of injury were excluded.

The primary exposure variable was mechanism of injury, which was categorized a priori as motorized road injury (automobiles, motorcycles, e-bikes, and other powered micromobility devices), non-motorized road injury (bicycles, push scooters, skateboards, and pedestrians not operating a powered device), or falls (including ground-level, stairs, ladder, and other fall-related injuries). Demographic variables included age, sex, and race or ethnicity. Additional variables abstracted from the registry included insurance payer type (commercial, Medicare, Medicaid, or self-pay/other), alcohol use at presentation, and the presence of orthopedic injuries or consultations. Measures of injury severity and physiology included emergency department GCS score and ISS. Resource utilization variables comprised ICU days, ventilator days, and total hospital length of stay (LOS). Mortality was ascertained from in-hospital death records.

Primary outcomes were mortality, ICU utilization, ventilator use, and hospital LOS. Secondary outcomes included TBI severity category (mild, moderate, or severe based on GCS) and the presence of orthopedic injury. Continuous variables were summarized as means with standard deviations (SD) and medians with interquartile ranges (IQR), while categorical variables were reported as counts and percentages. Group comparisons across the three injury mechanisms were performed using chi-square or Fisher’s exact tests for categorical variables and Kruskal–Wallis tests for continuous variables. As a prespecified secondary analysis, we conducted pairwise comparisons between motorized and non-motorized road injuries using Wilcoxon rank-sum tests for continuous variables (ISS, GCS, ICU days, ventilator days, LOS) and chi-square tests for categorical variables (alcohol use, sex, minority status, and ventilator use). All tests were two-sided with a significance threshold of α = 0.05. Missing data were minimal, and analyses were conducted using pairwise deletion. Statistical analyses were performed using SAS software (SAS Institute Inc., Cary, NC).

## Results

### Study population and mechanism distribution

Among 1,131 trauma patients with documented TBI falls accounted for the majority of cases, (n=1,020; 90.2%), followed by non-motorized road injuries (n=62; 5.5%) and motorized road injuries (n=49; 4.3%) (Table 1). Mechanism frequencies differed significantly across demographic and clinical characteristics.

**Table 1.**
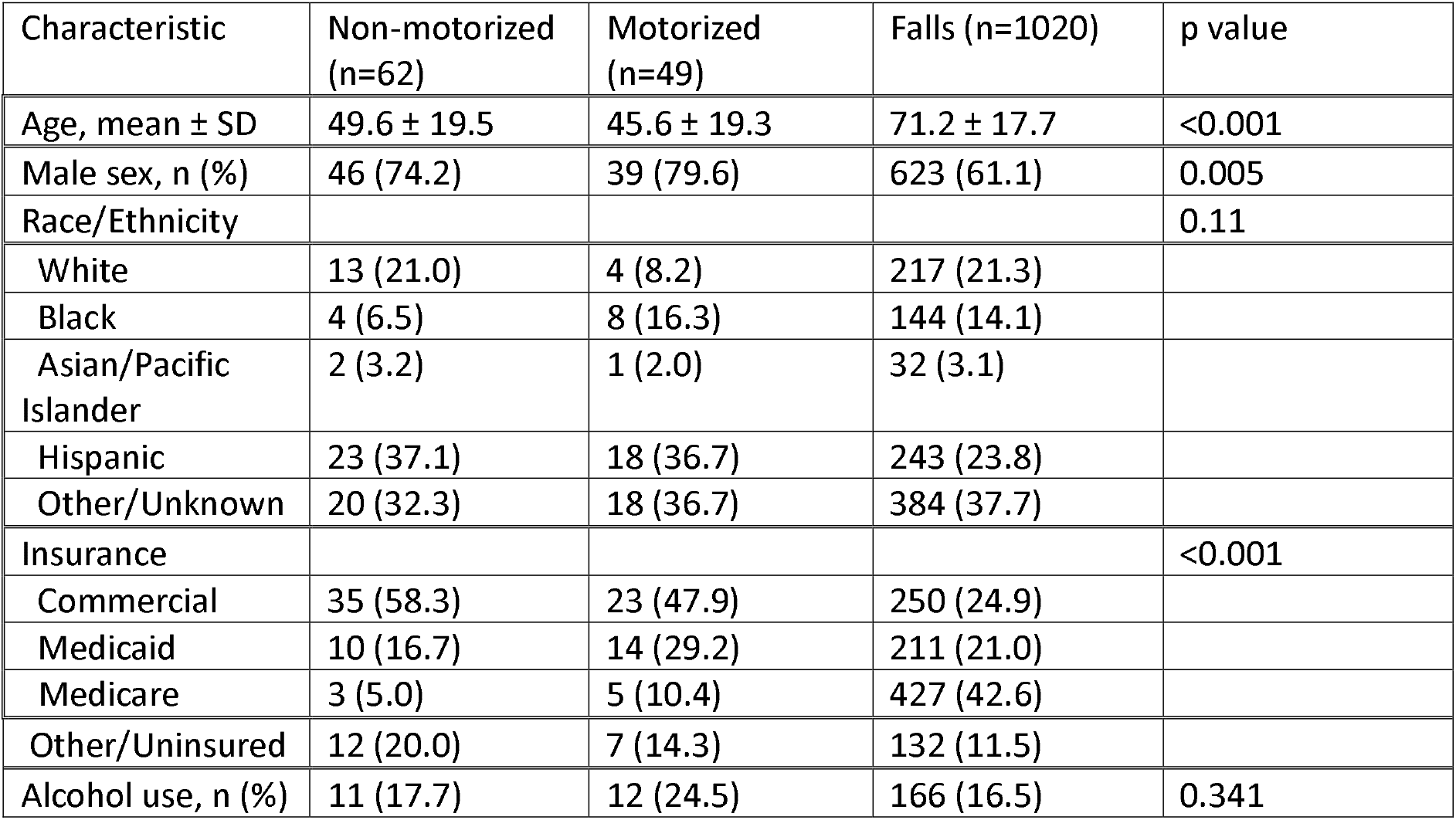
Demographic characteristics by mechanism of injury.

### Demographic Characteristics

Patients with fall-related TBI were substantially older than those with road related injuries (mean/median age 71.2/74.5 years vs 49.6/49 for non-motorized and 45.6/41.7 for motorized). The proportion of males were significantly higher in both non-motorized (74.2%) and motorized (79.6%) injuries compared with falls (61.1%) (p=0.00050)

Race and ethnicity also varied by mechanism. Hispanic patients constituted a markedly larger proportion of non-motorized (37.1%) and motorized (36.7%) injuries compared with falls (23.8%). In contrast, White patients accounted for 21.3% of fall-related TBIs, 21.0% of non-motorized injuries, and only 8.2% of motorized injuries. Black patients represented 14.1% of falls, 6.5% of non-motorized, and 16.3% of motorized injuries.

Insurance status reflected the age distribution: Medicare predominated among falls (42.6%), whereas commercial insurance was more common among non-motorized (58.3%) and motorized (47.9%) injuries. Medicaid coverage was highest among motorized injuries (29.2%). Alcohol use at presentation did not differ significantly across mechanisms (p=0.3408).

### Injury severity and clinical course

Measures of injury severity demonstrated significant variation by mechanism. (Table 2) Mean ISS was highest among motorized injuries (15.1), followed by falls (14.1) and non-motorized injuries (12.0) (p=0.04). Arrival Glasgow Coma Scale (GCS) scores were lowest in motorized injuries (mean 12.6) compared with non-motorized (13.7) and falls (13.8) (p=0.06).

**Table 2.**
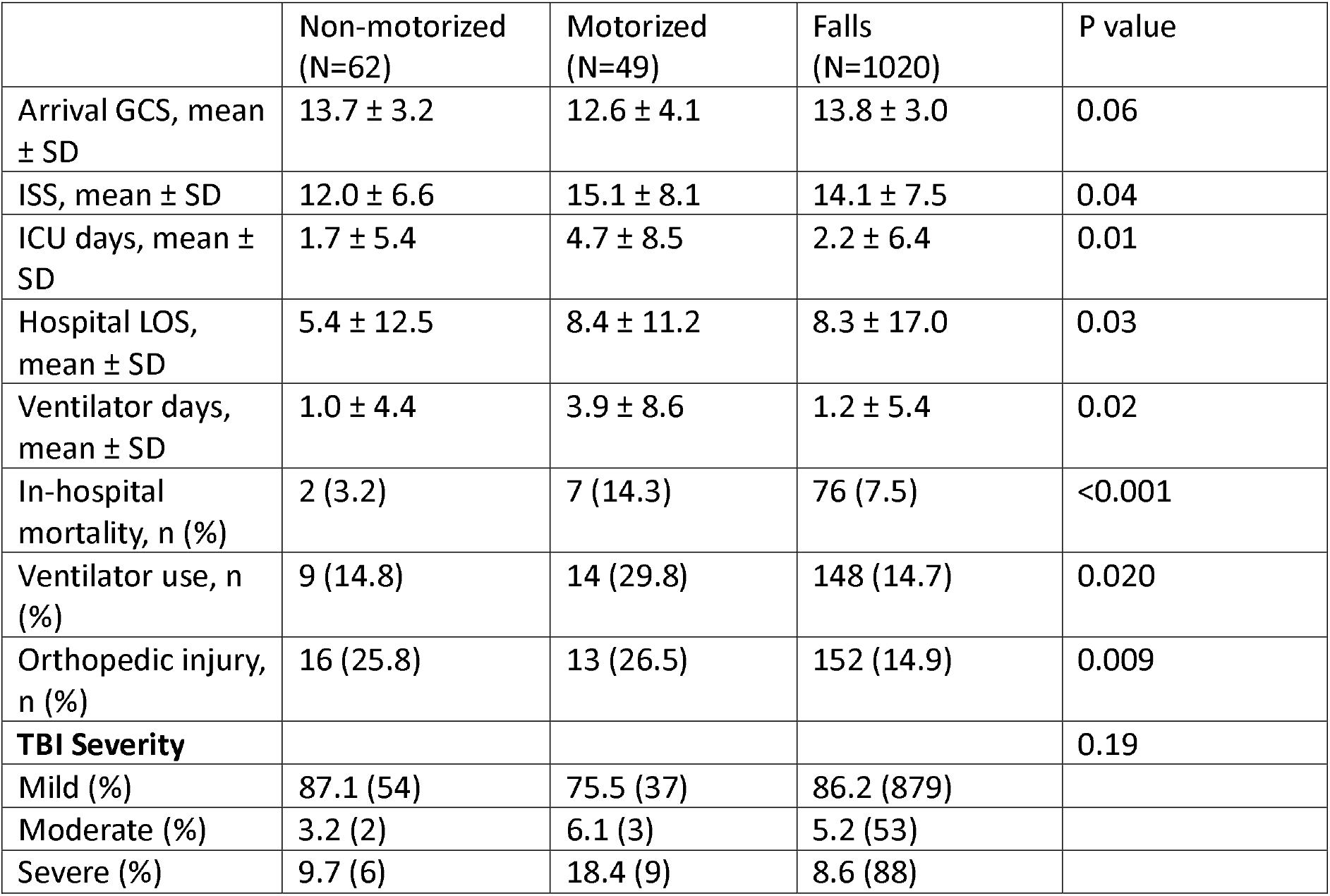
Outcomes and severity of non-motorized, motorized, and fall-related TBI.

Resource utilization differed significantly. Motorized injuries had the longest ICU stay (mean 4.7 days), compared with falls (2.2 days) and non-motorized injuries (1.7 days) (p=0.01). Motorized injuries also required the most ventilator support (mean 3.9 days), compared with non-motorized (1.0 days) and falls (1.2 days) (p=0.02). Hospital LOS was longest in motorized injuries (mean 8.37 days), similar to falls (8.30 days), and higher than non-motorized injuries (5.39 days) (p=0.03). Mechanical ventilation use differed significantly across groups (29.8% motorized, 14.8% non-motorized, 14.7% falls; p=0.02). Orthopedic injuries were more common in road injuries (25.8% non-motorized, 26.5% motorized) than in falls (14.9%) (p=0.009).

TBI severity classification also demonstrated trends across mechanisms. Moderate and severe TBIs were most frequent among motorized injuries (6.1% and 18.4%), compared with non-motorized injuries (3.3% and 9.7%) and falls (5.2% and 8.6%), although differences were not statistically significant (p=0.19).

### Mortality

Crude in-hospital mortality was 7.5% (85/1,131). Mortality differed by mechanism, highest among motorized injuries (14.3%), followed by falls (7.5%) and non-motorized injuries (3.2%). Deaths were disproportionately associated with falls (89.4% of all deaths), reflecting their predominance in the cohort (p<0.001 for distribution).

#### Pairwise Comparison of Motorized vs Non-Motorized Road Injuries

In the prespecified comparison limited to road injuries (n=111), motorized mechanisms demonstrated consistently greater injury severity and resource utilization than non-motorized mechanisms.(Table 3) Patients with motorized injuries had significantly higher ISS values (Z = 2.19, p = 0.014) and required more intensive care than patients with non-motorized injuries, reflected by longer ICU stays (Z = 2.16, p = 0.015) and more ventilator days (Z = 2.03, p = 0.021). Motorized injuries were also associated with a significantly longer hospital length of stay (Z = 2.13, p = 0.017). Arrival GCS tended to be lower among motorized injuries, although this difference did not reach statistical significance (Z = –1.57, p = 0.058). There were no significant differences between motorized and non-motorized injuries in age (p = 0.12), sex (p = 0.505), minority status (p = 0.384), alcohol use (p = 0.384), or the binary indicator of ventilator use (p = 0.059).

**Table 3.**
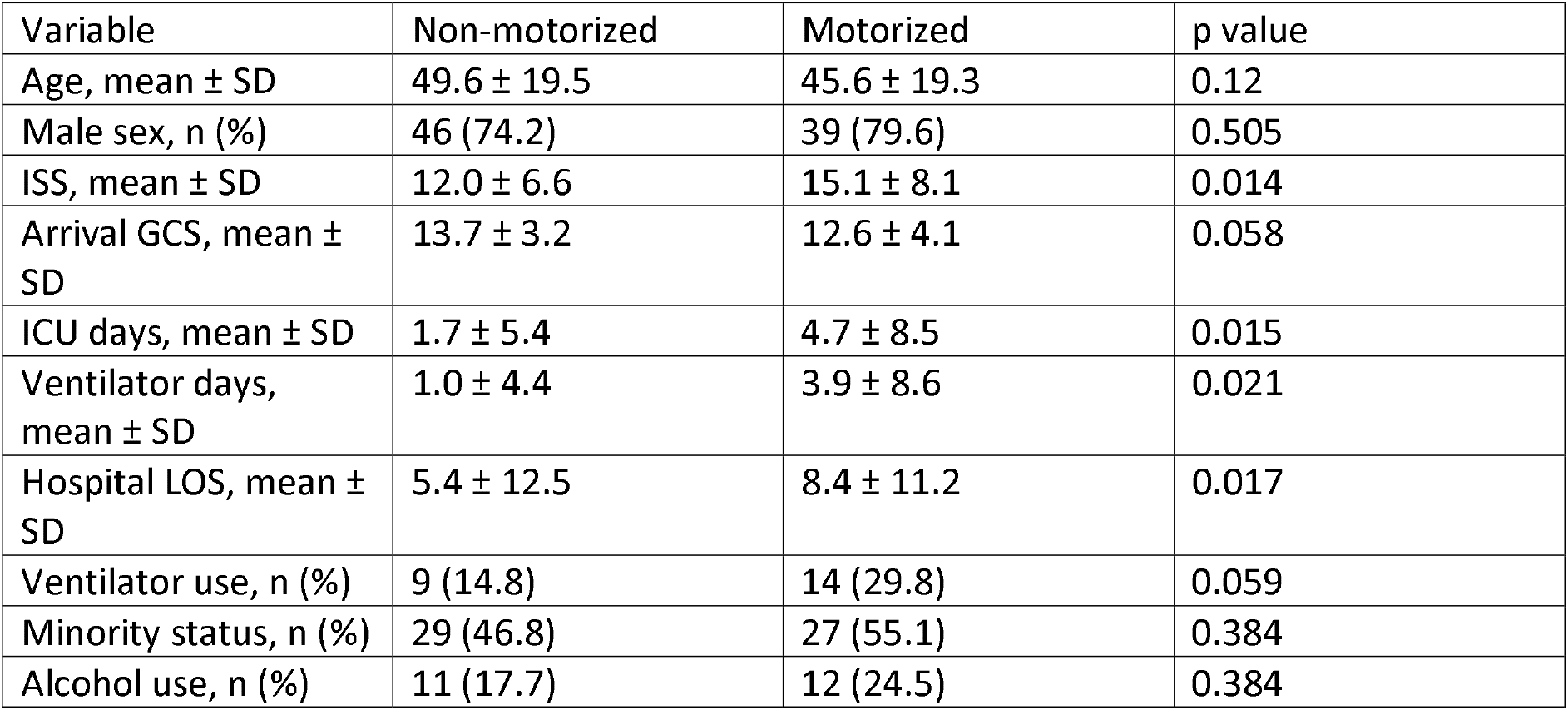
Pairwise Comparison of Road-Related TBIs.

## Discussion

In this single-center cohort of more than 1,100 TBI patients, falls accounted for the overwhelming majority of injuries, reflecting contemporary epidemiologic patterns in high-income countries and the growing burden of fall-related TBI among older adults.^6-8^ In contrast, motorized road injuries represented a small fraction of cases yet were associated with disproportionately greater injury severity, resource utilization, and in-hospital mortality when compared to non-motorized road injuries. These findings underscore the substantial heterogeneity of TBI burden by mechanism and highlight how relatively infrequent, high energy mechanisms contribute disproportionately to severe outcomes.

Clear demographic differences emerged across mechanisms. Fall-related TBI predominantly affected older adults and was strongly associated with Medicare coverage, consistent with age-related fall risk and medical vulnerability. Road-related injuries occurred in substantially younger patients and were more common among males, aligning with established patterns of vehicular and micromobility trauma.^1-8^ Notably, Hispanic patients constituted a significantly larger proportion of both motorized and non-motorized road injuries compared with falls. While this study was not designed to assess causality, this pattern may reflect differential exposure to urban transportation modes, occupational travel, or neighborhood-level infrastructure factors, and warrants further investigation in multicenter and population-based studies.

Motorized road injuries were consistently associated with greater injury severity than non-motorized mechanisms. Patients injured by motorized vehicles had higher ISS, longer ICU and ventilator durations, and longer hospital stays, despite only modest differences in arrival GCS. This finding suggests that motorized mechanisms produce greater multisystem trauma beyond neurologic injury alone, likely due to higher energy transfer and associated extracranial injuries. The significantly higher frequency of orthopedic injuries among road-related TBIs further supports this interpretation and is consistent with prior work linking polytrauma to increased resource utilization and prolonged hospitalization after TBI.^20-21^

The elevated crude mortality observed among motorized injuries (14%) reinforces the clinical impact of these high-energy mechanisms. Although fall-related TBI occurred in an older and more medically complex population, mortality among falls was lower than in motorized injuries, likely reflecting lower kinetic energy but higher baseline vulnerability. Together, these findings illustrate how injury mechanism interacts with patient age and physiologic reserve to shape outcomes, rather than acting as an isolated determinant of prognosis.

Our results parallel findings from international cohorts including the *International Traumatic Coma Project* and European trauma registries, which have reported higher ISS and greater resource utilization among motorized injuries compared with non-motorized or fall related mechanisms.^9-12^ However, most prior studies were conducted outside the United States, where transportation patterns, micromobility adoption, and healthcare access differ. By demonstrating similar trends in a large, diverse urban U.S. population, this study adds important domestic evidence to a limited literature and highlights the relevance of mechanism-specific risk in contemporary American trauma systems.

Alcohol use at presentation was common across mechanisms, but did not differ significantly between groups. This finding is consistent with prior registry based studies suggesting that while intoxication is prevalent in road related trauma, it may not independently predict injury severity or neurologic presentation once mechanism and other factors are considered.^18-19^ The lack of significant difference in our cohort may also reflect urban variability in alcohol screening or reporting thresholds. In contrast, orthopedic injury showed a strong association with road mechanisms, reinforcing its role as a marker of higher overall trauma burden.

Several limitations merit consideration. This was a single-center, retrospective analysis, limiting generalizability and precluding causal inference. Mechanism classification relied on registry abstraction and may not fully capture mixed or complex injury scenarios. Data on helmet use, vehicle speed, crash dynamics, and prehospital care were unavailable, and outcomes were limited to the in-hospital course. Residual confounding by comorbidities or injury context is therefore possible. Nonetheless, the use of standardized trauma registry variables strengthens internal validity and allows meaningful comparisons across mechanisms.

Despite these limitations, this study provides clinically relevant insight into how injury mechanism shapes TBI severity and outcomes in an urban U.S. trauma population. While falls dominate by frequency, motorized road injuries account for a disproportionate share of severe cases, intensive care utilization, and in-hospital mortality.

## Conclusion

In this large, single-center cohort of patients with diagnosed traumatic brain injury, falls accounted for most cases and primarily affected older adults. However, motorized road injuries—though far less frequent—were associated with greater injury severity, increased ICU and ventilator use, longer hospital stays, and higher mortality compared with both non-motorized road injuries and falls. The overrepresentation of Hispanic patients among road-related TBIs further highlights potential disparities in exposure and risk within urban environments.

These findings emphasize that injury mechanism remains a critical determinant of trauma burden and outcome. Effective TBI prevention must therefore be mechanism-specific, combining fall-prevention strategies for older adults with targeted interventions for road-related injuries, including helmet use, safer micromobility infrastructure, and enforcement of motor vehicle safety and alcohol-use policies. Future multicenter studies should examine long-term functional outcomes and social determinants of risk to better inform prevention and post-acute care strategies.

## Data Availability

All data in the present study are available upon reasonable request to the Mount Sinai Health System, Department of Surgery and completion of an IRB application for use of the data.

## Notes

### Competing Interest Statement

The authors have declared no competing interest.

### Funding Statement

This study did not receive any funding.

### Author Declarations

The institutional review board of the Mount Sinai Health System (Icahn School of Medicine at Mount Sinai) gave ethical approval for this work as exempted research utilizing de-identified patient data.

